# Robotically-controlled three-dimensional micro-ultrasound for prostate biopsy guidance

**DOI:** 10.1101/2022.12.23.22283894

**Authors:** Reid Vassallo, Tajwar Abrar Aleef, Qi Zeng, Brian Wodlinger, Peter Black, Septimiu E. Salcudean

**Affiliations:** School of Biomedical Engineering, The University of British Columbia, 251-2222 Health Sciences Mall, Vancouver, V6T 1Z3, BC, Canada; MD/PhD Program, The University of British Columbia, 317-2194 Health Sciences Mall, Vancouver, V6T 1Z3, BC, Canada; Department of Electrical and Computer Engineering, The University of British Columbia, 5500-2332 Main Mall, Vancouver, V6T 1Z4, BC, Canada; Exact Imaging, 15-7676 Woodbine Avenue, Markham, L3R 2N2, ON, Canada; Department of Urologic Sciences, The University of British Columbia, 2775 Laurel Street, Vancouver, V5Z 1M9, BC, Canada

**Keywords:** Micro-ultrasound, prostate cancer, three dimensional, robotics

## Abstract

**Purpose:** Prostate imaging to guide biopsy remains unsatisfactory, with current solutions suffering from high complexity and poor accuracy and reliability. One novel entrant into this field is microultrasound (microUS), which uses a high frequency imaging probe to achieve very high spatial resolution, and achieves prostate cancer detection rates equivalent to multiparametric magnetic resonance imaging (mpMRI). However, the ExactVu transrectal microUS probe has a unique geometry that makes it challenging to acquire controlled, repeatable three-dimensional (3D) transrectal ultrasound (TRUS) volumes. We describe the design, fabrication, and validation of a 3D acquisition system that allows for the accurate use of the ExactVu microUS device for volumetric prostate imaging.

**Methods:** The design uses a motorized, computer-controlled brachytherapy stepper to rotate the ExactVu transducer about its axis. We carry geometric validation using a phantom with known dimensions and we compare performance with magnetic resonance imaging (MRI) using a commercial quality assurance anthropomorphic prostate phantom.

**Results:** Our geometric validation shows accuracy of 1 mm or less in all three directions, and images of an anthropomorphic phantom qualitatively match those acquired using MRI and show good agreement quantitatively.

**Conclusion:** We describe the first system to acquire robotically-controlled 3D microUS images using the ExactVu microUS system. The reconstructed 3D microUS images are accurate, which will allow for applications of the ExactVu microUS system in prostate specimen and *in vivo* imaging.

## 1 Introduction

Prostate cancer (PCa) is the second-most frequently diagnosed cancer in males worldwide, and the World Health Organization (WHO) estimates that 2020 saw over 1.4 million new diagnoses of the disease, resulting in over 375,000 deaths [1]. The standard diagnostic method for PCa is transrectal ultrasound (TRUS)-guided biopsy, where a series of tissue samples are obtained using an 18 gauge needle to determine a histological diagnosis [2, 3]. However, suspicious prostate lesions cannot be reliably targeted because they often appear isoechoic on standard TRUS B-mode images [4]. This has lead to a standardized biopsy method being employed, where a series of approximately 12 regularly-spaced tissue samples are obtained, and the primary purpose of the TRUS image is to visualize the external border of the prostate [5]. This standardized biopsy technique, however, leads to a false negative rate of over 30% [6], because it is unclear if a negative result is due to not sampling the correct area of the prostate (representing a false negative result), or if no cancerous cells are present (representing a true negative result). Although repeat biopsy is an option after a negative result, this carries its own risk as up to 6.3% of patients require hospitalization due to complications following prostate biopsy [7].

Being able to perform targeted biopsy on suspicious lesions would address these limitations, and this can currently be done using multiparametric magnetic resonance imaging (mpMRI) [8]. However, this approach suffers from limitations which are inherent to MRI, such as high cost, low accessibility, and the inability to acquire images in real-time. Another current area of research is using multiparametric ultrasound (mpUS) to visualize areas which are suspicious for prostate cancer [9], but it has not yet demonstrated equivalency to mpMRI for this purpose [10, 11].

A recent addition to the prostate imaging arsenal is micro-ultrasound (microUS), particularly by using the ExactVu imaging system (Exact Imaging, Markham, Canada). This system uses a side-fire ultrasound (US) probe that can reach 29 MHz to image the prostate (compared to the usual 9-12 MHz [12]), acquiring images with spatial resolution of 70 *μ*m (which is approximately the size of prostatic ducts [13]). Meta-analyses have demonstrated that microUS is non-inferior to mpMRI for targeted prostate biopsy [14, 15], and an upcoming trial will compare mpMRI fusion biopsy to targeted biopsy with microUS alone [16].

One limitation of the ExactVu system is that it can only acquire two dimensional (2D) images natively, and its unique probe geometry does not allow for the easy use of mechanical sweep systems which have been developed for other TRUS probes. As can be appreciated in Figure 1, most TRUS probes are essentially cylindrical in shape, so that rotating the handle of the probe about its principal axis will also rotate the element array about that same axis, creating a 3D image with a well-defined geometry. The ExactVu microUS probe, on the other hand, does not have parallel axes between the handle of the probe and the element array, so rotating about the handle’s principal axis will result in an incorrect 3D image with incorrect spatial geometry.

**Fig. 1.**
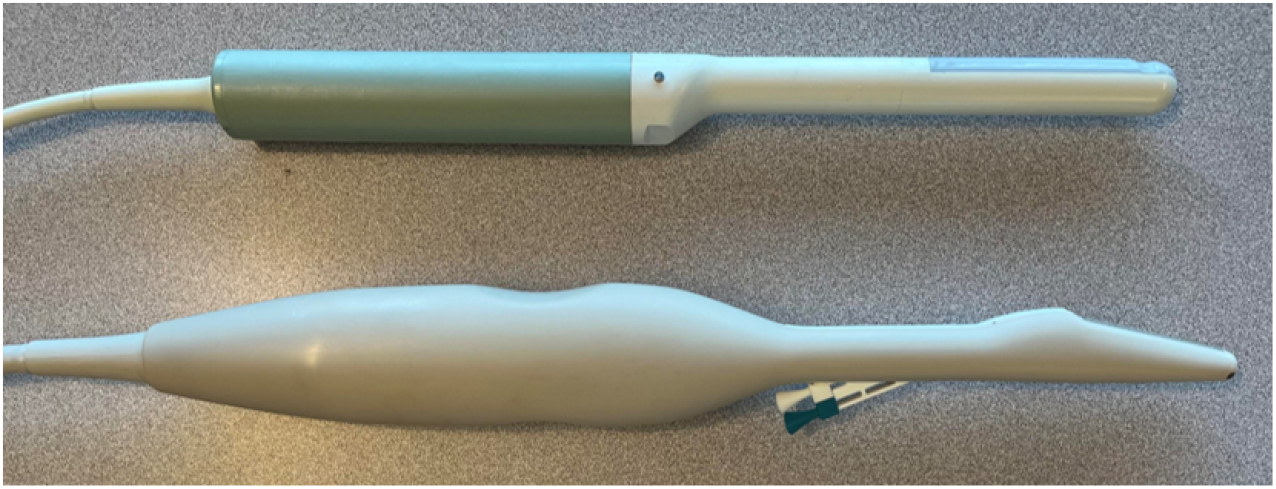
An image showing the ExactVu microUS probe (bottom) compared to a traditional side-fire TRUS probe geometry (top). The traditional side-fire TRUS probe here is the E14CL4b endocavity biplane transducer (BK Medical, Herlev, Denmark).

The ability to acquire reliable 3D images using microUS will (i) allow for its inclusion in already-defined imaging workflows, such as 3D elastography [17], (ii) better facilitate MRI-to-US registration for fusion biopsies, (iii) allow for more accurate microUS-guided biopsies or brachytherapy without the necessity of MRI for volumetric information, and (iv) serve as an important tool in robotic surgery guidance [18].

The objective of this paper is the design and validation of a novel robotically-controlled system to generate accurate 3D images using the ExactVu microUS system.

## 2 Methods

### 2.1 System Design

We follow the design approach for the robotic TRUS system [19] designed for Ultrasonix and BK Medical systems. This will allow us to leverage upon existing infrastructure, which has been approved for previous *in vivo* studies [20, 21].

The overall system is designed to be integrated with the clinical CIVCO EX-II stepper (CIVCO, Coralville, IA, USA) by replacing its native encoder with an external motor (Faulhaber, Schönaich, Germany). This external motor is controlled by a motor control box which includes a microcontroller and can be programmed directly using the ExactVu system, which runs a Windows operating system in its research mode (Microsoft, Redmond, WA, USA), allowing the installation of our custom software to control the motor. Additionally, this motor is fitted with an optical encoder, ensuring accurate imaging increments.

To overcome the geometric challenges presented by the ExactVu probe’s shape, we designed an adapter to align the probe with the robot such that the axis of rotation for the robot is parallel with the lateral direction of the ultrasound imaging array. The precise shape of the probe was determined using a Artec Leo handheld 3D scanner (Artec 3D, Senningerberg, Luxembourg), which was post-processed and imported into the computer-aided design (CAD) software Solidworks (Dassault Systèmes, Vélizy-Villacoublay, France), where the adapter was created to align the axes based on the known angle between the center axis of the probe and the element array. The final design is shown in Figure 2.

**Fig. 2.**
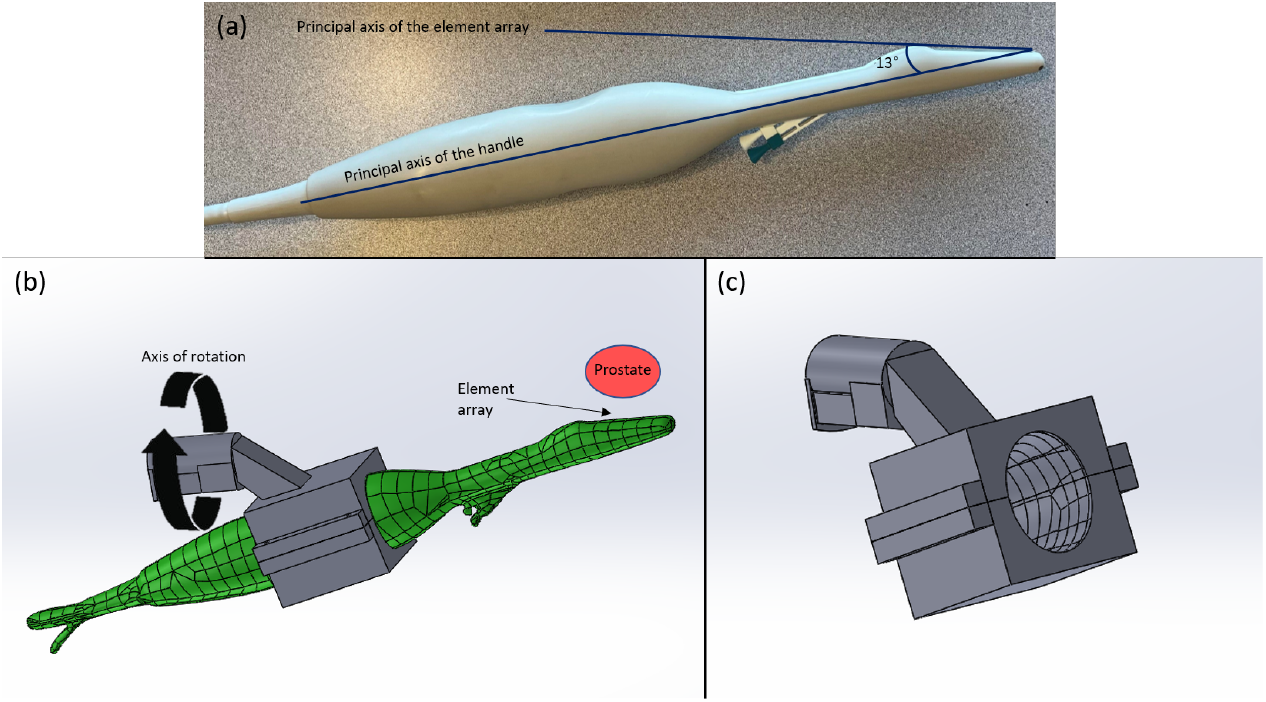
A figure describing the design of this system. (a) An image of the ExactVu microUS probe, with the principle axes of the handle and element array defined, as well as the angle between them. (b) A screen capture of the computer-aided design (CAD) model of the ExactVu probe created by 3D scanning (green) and the component to transform the axis of rotation. (c) A second view of the CAD model of our component without the microUS probe.

The device was created using an Afinia H800+ (Afinia, Chanhassen, MN, USA) fused deposition modeling (FDM) 3D printer with polylactic acid (PLA) filament, and post-processed to ensure accurate alignment with the existing robotic TRUS system.

There is a 13 degree angle between the lateral direction of the element array of the probe and the principal axis of the handle. This information was used to choose the angle of our device. The center axis of the element array was then offset to be 3 mm above the center of rotation of the robotic system, so that it would travel along a circular path with a radius of 3 mm. This allows the system to maintain contact with the imaging surface, and thus acoustic coupling.

The interface between our device and the probe was designed such that it affixes onto indentations in the handle of the probe, usually intended as an ergonomic grip. These indentations are on two sides of the device and are not identical. This design ensures that the probe can only be attached in one orientation and keeps it securely in place without requiring any adhesives or other modifications to the probe.

Scan conversion was performed in MATLAB R2022b (The Mathworks, Natick, MA, USA) to combine the series of acquired 2D B-mode images into a single 3D volume using linear interpolation, with 0.2 mm isotropic pixel spacing. The increment between acquired 2D images was 1 degree.

### 2.2 System Validation

Our system was validated using several methods, to ensure it created accurate 3D B-mode volumes. First, it underwent geometric validation in all three principal directions and then it was compared against a 3.0 T MRI scanner by imaging a commercial quality assurance prostate phantom, providing qualitative and quantitative results.

#### 2.2.1 Geometric Validation

Geometric validation of this imaging system was performed using a 3D printed fCal 2.1 phantom [22], which was strung with 20 *μ*m diameter tungsten wire, such that crossings were present with a known distance between each of them, allowing comparison between this known distance and what is measured in the reconstructed 3D microUS image. Such fine wire is required due to the very precise in-plane spatial resolution of the ExactVu system, as described above. This phantom was imaged inside a water bath in several positions and orientations, so that measurements could be made at various points in the imaging volume to form representative results. These images were acquired with an image depth setting of 50 mm. Due to the high frequency of this device, the maximum image depth is lower than in other systems.

It is known from construction that wire crossings on the same row are 15 mm apart, while 5 mm separates rows, which can be seen in Figure 3. The 3D reconstructed images were analyzed in 3D Slicer [23], with wire crossing locations manually confirmed and segmented by reviewing all three reconstructed planes and the volume rendered image. Euclidean distances were calculated between relevant points, and the absolute difference between this distance and the value known from construction (15 mm or 5 mm) represents our geometric validation error measurement.

**Fig. 3.**
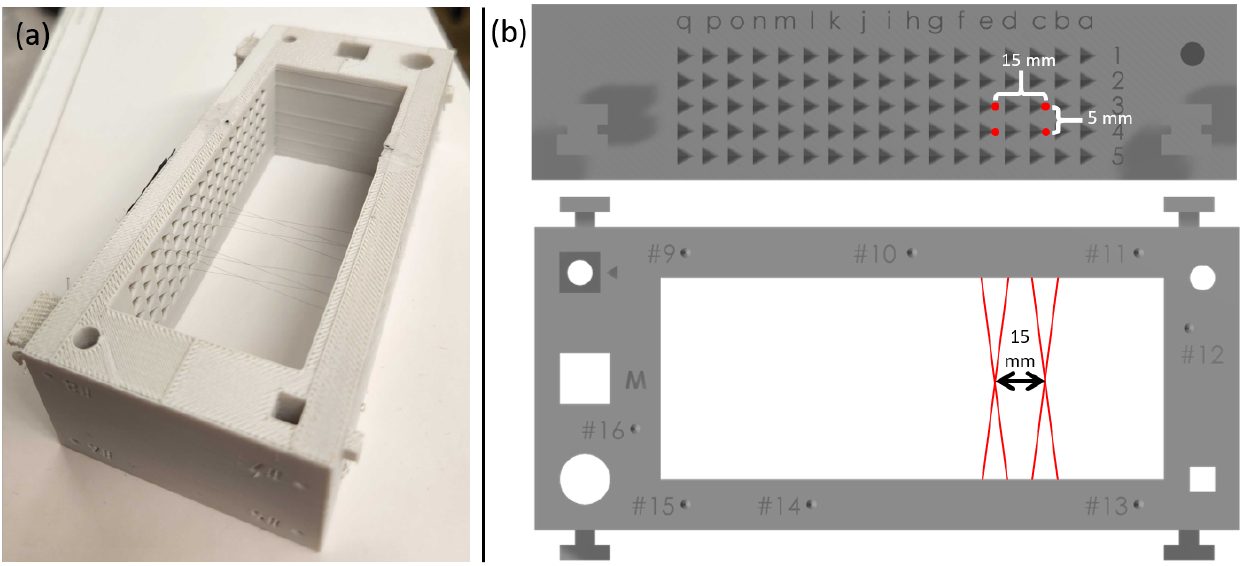
An image showing the fCal phantom with crossings of 20 *μ*m wire. The distance between crossings is known to be 15 mm within the same row, and 5 mm between rows by construction. (a) A photograph of the phantom. (b) Schematics showing the fCal phantom, the wire crossings and the distance between them in two projections.

A cartoon depiction of the approximate measurement locations is shown in Figure 4 in two orthogonal projections.

**Fig. 4.**
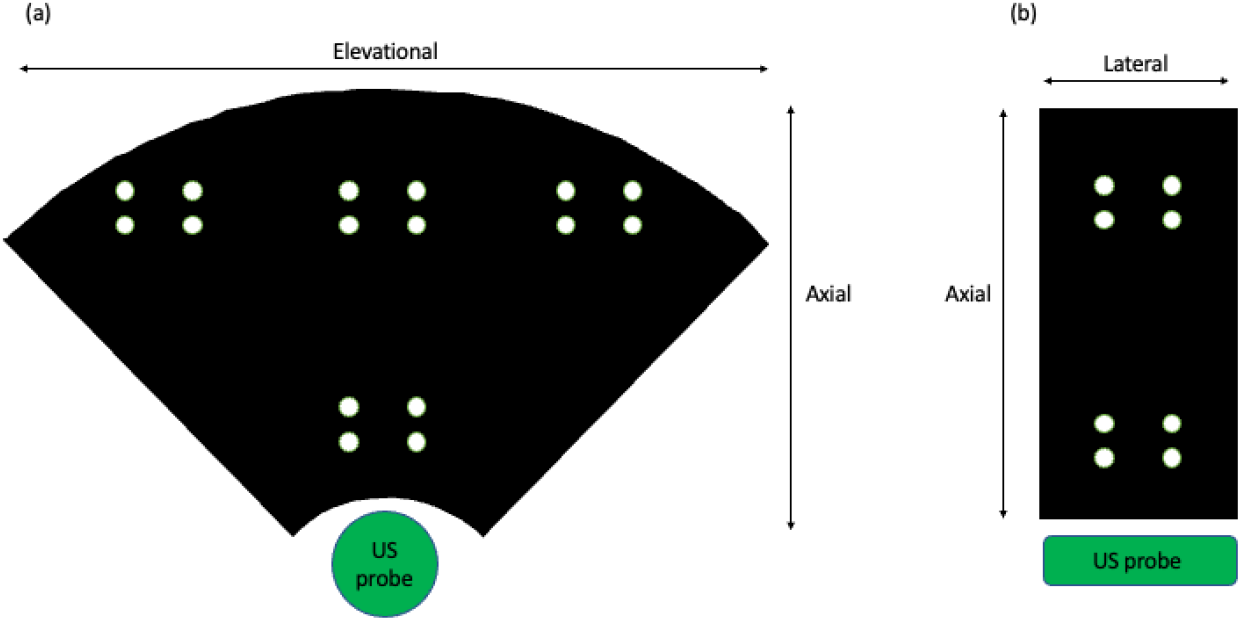
A cartoon representation of the approximate measurement locations in projections of the image volume, showing (a) the elevational and axial directions, and (b) the lateral and axial directions.

For the purposes of this paper, the lateral, axial, and elevational directions will be used in reference to our image volume, and will be defined with respect to the image plane in the center of our volume. These directions are also shown in Figure 4.

#### 2.2.2 Phantom Validation

Images were acquired of a commercial quality assurance prostate imaging phantom using our 3D microUS system, and compared to images of the same phantom acquired with MRI.

The phantom is a Tissue Equivalent Ultrasound Prostate Phantom 053L (CIRS, Norfolk, VA, USA), which is intended for use with side-fire TRUS probes. This phantom includes several simulated anatomical features, including the rectal wall, seminal vesicles, urethra, and three simulated spherical lesions (each approximately 10 mm in diameter) inside the prostate. The dimensions of the prostate in this phantom are 5 × 4.5 × 4 cm, and the overall phantom dimensions are 11.5 × 7 × 9.5 cm.

The parameters of the microUS system were identical to those described above in the geometric validation procedure (50 mm image depth and images acquired every 1 degree, resulting in a volume with 0.2 mm isotropic voxel spacing).

The *T*_2_-weighted MRI image was acquired using a Philips Ingenia Elition 3T X (Philips Healthcare, Amsterdam, Netherlands), with voxel spacing of 1.5 × 1.5 × 1.5 mm. The relaxation time was 1800 ms, echo time was 80 ms, and flip angle was 90 degrees.

The 3D microUS and MRI image volumes were compared by first rigidly registering them using 3D Slicer, and then selecting representative images from all three reconstructed planes for qualitative comparison.

For a quantitative comparison of our reconstructed image, the lesions in the phantom were manually segmented in both volumes, and the distance between the centroids of these segmentations was calculated in each image volume. This measurement was then compared between the two image volumes.

## 3 Results

### 3.1 Geometric Validation

The results from our geometric validation are presented in Figure 5, which demonstrates the sub-millimeter accuracy of our reconstructed 3D microUS images in all three dimensions. These results are further broken down into measurements taken in the near-field of the image (approximately 15-20 mm from the probe), and those taken when the wire crossings are in the far-field (approximately 35-40 mm from the probe). Overall, the mean geometric validation errors are 0.53 mm, 0.17 mm, and 0.30 mm in the elevational, lateral, and axial directions, respectively.

**Fig. 5.**
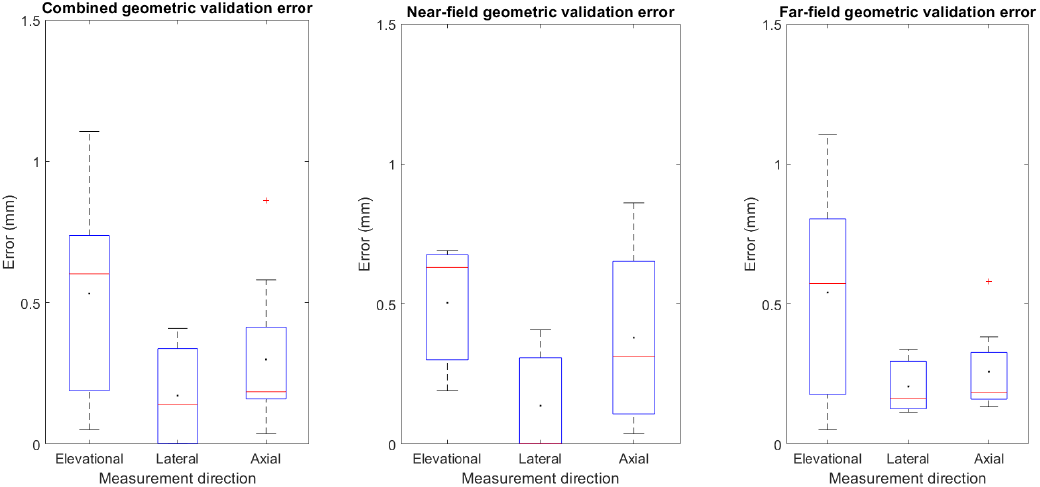
A boxplot of the of the measured errors from our 3D microUS images of an fCal phantom whose wire crossings are known distance from each other by construction, showing the median and and quartile values, with the mean values overlaid in black. The left plot shows all measurements combined, while the middle plot shows error measurements when the crossings are in the near-field (approximately 15-20 mm from the probe), and the right plot shows these measurements in the far-field (approximately 35-40 mm from the probe).

### 3.2 Phantom Validation

Representative images of the prostate phantom from all three planes (transverse, sagittal, and coronal) of our reconstructed 3D microUS volume are compared to that of *T*_2_-weighted MRI after rigid registration in Figure 6, demonstrating very good agreement.

**Fig. 6.**
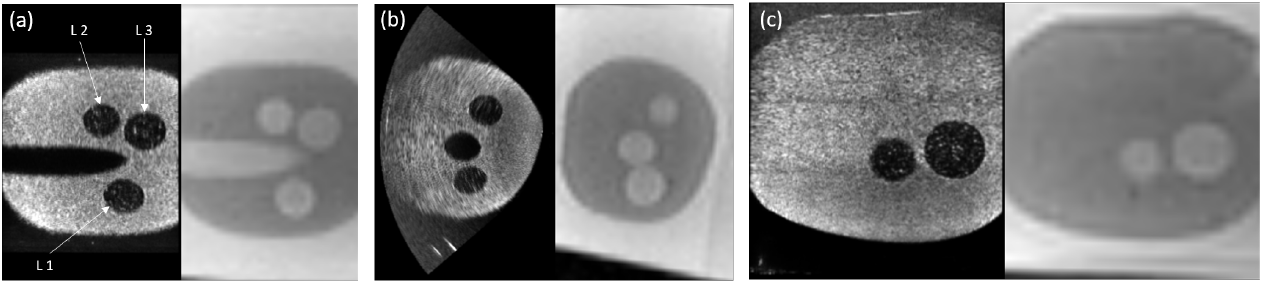
A comparison between our reconstructed 3D microUS volume and *T*_2_-weighted MRI images of the same prostate phantom after rigid registration, in (a) the coronal, (b) the transverse, and (c) the sagittal planes. (a) The lesions are each labelled as L1, L2, and L3, and these are used for the quantitative results in Table 1.

The quantitative results of the comparison between the microUS and MRI volumes are shown in Table 1. The mean difference of the measurements between lesion centroids was 1.09 mm.

**Table 1.** Quantiative results of microUS-to-MRI comparison

|  | MRI | MicroUS | Difference |
| --- | --- | --- | --- |
| L1-L2 | 21.24 mm | 19.84 mm | 1.51 mm |
| L1-L3 | 18.58 mm | 17.09 mm | 1.40 mm |
| L2-L3 | 11.41 mm | 11.05 mm | 0.37 mm |

## 4 Discussion

The aim of this paper was to describe the design and validation of a novel method to acquire 3D microUS images using the ExactVu system, for use in prostate cancer biopsy guidance. This system was validated using a wire phantom, with distances between wires known from construction showing submillimeter error in all three reconstructed directions. There is anisotropy to the amount of error in the three directions, which is to be expected due to the differing amount of interpolation that must be performed in each of the directions. Although our geometric validation results show some error, they compare favourably to previously-published standard values for US systems of 1.5 mm error in the in-plane vertical and horizontal directions (corresponding to axial and lateral here) [24, 25], and 2-3 mm in the elevational direction [24]. Results of the phantom validation demonstrate good agreement between our reconstructed volume and a volume of the same phantom acquired using MRI, signifying that this system can lead to accurate imaging of anatomy at very high resolutions in three dimensions.

The reconstructed image from our 3D microUS system provides an accurate representation of the volume being imaged, shown by the millimeter scale or better accuracy in distance measurements, as well as its favourable comparison to an MRI image of the same prostate phantom. This will provide excellent anatomical information of the prostate in high resolution and three dimensions.

This development method can be easily repeated for any TRUS system to acquire reliable 3D US volumetric imaging, provided the geometry of the probe is known and well-defined, namely the angle between the principal axes of the handle and element array and the offset between them.

### 4.1 Limitations

This study represents the initial validation and characterization of this 3D microUS system. Further refinements and validation will likely be required before it can be used clinically.

There were also potential sources of error in the construction and validation of this system. Namely, there are errors associated with the 3D scanner and 3D printer used here. Error from the 3D printer will be addressed in the future by fabricating the next generation of this system using more advanced techniques, such as a sterolithography 3D printer or precision subtractive manufacturing techniques.

Some of the distance measurement error can also likely be accounted for by the differences in speed of sound between soft tissue and water, as argued in [26], or the fact that the wire crossing points were manually determined, providing a source for human error.

Our quantitative phantom validation results are likely impacted by the relatively poor 1.5 mm isotropic voxel spacing of our MRI volume, which would magnify any lesion centroid localization error arising from the manual segmentation.

### 4.2 Future Work

This work represents a necessary step for extending the abilities of the ExactVu microUS system. The ability to image in 3D will allow for the implementation of cutting-edge 3D elastography methods [17], improved mpUS methods for prostate cancer diagnosis and biopsy guidance [27], or robotic surgery guidance [18].

## 5 Conclusion

This paper presents the design, construction, and validation of the first known system to capture accurate robotically-controlled 3D microUS images with the ExactVu system. Error was measured in all three prinicipal directions of the image, showing millimeter scale or better accuracy, and phantom images compared favourably to MRI. Future work will include using this development as a springboard to extend the utility of this microUS system, such as by developing a 3D microUS elastography system to leverage microUS’ inherent advantages over traditional US, including superior spatial resolution.

## Data Availability

All data produced in the present study are available upon reasonable request to the authors

## 5.1 Acknowledgements

This work was funded with a Natural Sciences and Engineering Research Council (NSERC) Canada Graduate Scholarship - Doctoral (CGS-D) and the C.A. Laszlo Chair held by Professor Salcudean.

